# The human host response to monkeypox infection: a proteomic case series study

**DOI:** 10.1101/2022.07.27.22278027

**Authors:** Ziyue Wang, Pinkus Tober-Lau, Vadim Farztdinov, Oliver Lemke, Torsten Schwecke, Sarah Steinbrecher, Julia Muenzner, Helene Kriedemann, Leif Erik Sander, Johannes Hartl, Michael Mülleder, Markus Ralser, Florian Kurth

**Author notes:** These authors contributed equally (Ziyue Wang, Pinkus Tober-Lau, Vadim Farztdinov, Michael Mülleder, Markus Ralser, Florian Kurth).

## Abstract

Monkeypox (MPX) is caused by the homonymous orthopoxvirus (MPXV) known since the 1970s to occur at low frequency in West and Central Africa. Recently, the disease has been spreading quickly in Europe and the US. The rapid rise of MPX cases outside previously endemic areas and the different clinical presentation prompt for a better understanding of the disease, including the development of clinical tests for rapid diagnosis and monitoring. Here, using Zeno SWATH MS - a latest-generation proteomic technology - we studied the plasma proteome of a group of MPX patients with a similar infection history and clinical severity typical for the current outbreak. Moreover, we compared their proteomes to those of healthy volunteers and COVID-19 patients. We report that MPX is associated with a strong and characteristic plasma proteomic response and describe MPXV infection biomarkers among nutritional and acute phase response proteins. Moreover, we report a correlation between plasma protein markers and disease severity, approximated by the degree of skin manifestation. Contrasting the MPX host response with that of COVID-19, we find a range of similarities, but also important differences. For instance, Complement factor H-related protein 1 (CFHR1) is induced in COVID-19, but suppressed in MPX, reflecting the different role of the complement system in the two infectious diseases. However, the partial overlap between MPX and COVID-19 host response proteins allowed us to explore the repurposing of a clinically applicable COVID-19 biomarker panel assay, resulting in the successful classification of MPX patients. Hence, our results provide a first proteomic characterization of the MPX human host response based on a case series. The results obtained highlight that proteomics is a promising technology for the timely identification of disease biomarkers in studies with moderate cohorts, and we reveal a thus far untapped potential for accelerating the response to disease outbreaks through the repurposing of multiplex biomarker assays.

## Introduction

The outbreak of monkeypox (MPX) with currently more than 15,000 confirmed infections worldwide is exceptional in scale and spread [1], and has recently been declared a global emergency by the WHO [2]. MPX is caused by the zoonotic monkeypox virus (MPXV), a member of the genus *Orthopoxvirus* [3]. The first human MPX case was reported in 1970 in the Democratic Republic of the Congo (DRC), which is still the region with the highest level of endemicity in Africa [4]. Several outbreaks have been reported from African countries during the past decades, but research on MPX has largely been neglected. The clinical presentation often includes typical skin lesions, fever, and swollen lymph nodes. MPX is usually self-limiting, but severe cases can occur and a case fatality rate of 1–10% has been reported from Africa, with generally higher case fatality associated with the Central African compared to the West African virus clade [4].

The molecular epidemiology of the current MPX outbreak suggests that the current strain is closely related to that of a 2018–2019 outbreak in the United Kingdom and may have been circulating in the human population for some time, possibly with adaptation to the human host [5,6]. In the current outbreak, there is a clear predominance of infections among men who have sex with men (MSM), and several large public events have been associated with rapid emergence of cases in different parts of the world. Currently, transmission via close skin and mucosal contact, possibly including sexual transmission, seems likely [7–10]. Even though the current outbreak is still in its early stages, a self-limiting course cannot be assumed; rather, it is a longer-term public-health problem that will hopefully bring diagnostic and therapeutic benefits to endemic African countries.

The COVID-19 pandemic has reminded us of the need to create infrastructure and methodologies to respond rapidly to emerging pathogens. Mass-spectrometry-based proteomics is one of the emerging technologies in this regard, which due to the technical and analytical advances during the last years is increasingly moving into clinical application [11–14]. In the early phase of the COVID-19 pandemic, proteomic analyses provided rapid insights into the nature of the human response to SARS-CoV-2 and captured hallmarks of its immune evasion strategies and pathophysiology, including its impact on the complement system, coagulation cascade, and inflammatory and nutritional response machinery [12,15–19]. Furthermore, proteomic signatures turned out to accurately classify disease severity in COVID-19 and allow for outcome prediction weeks in advance [19–21]. Recently, we were able to show the strength of mass-spectrometry-based proteomics for rapid translation to medical care by generating a routine-applicable proteomic biomarker panel which predicted COVID-19 severity and outcome in a multi cohort study [22]. While such proteomic assays are currently primarily used to monitor clinical trials, they are increasingly being considered for their potential to optimize treatment and resource allocation, as well as to aid navigation of difficult triaging situations in the event of a pandemic.

Here, we describe the proteomic changes in a case series, a group of patients hospitalized due to MPXV infection that share a similar disease and infection history. We detect significant and consistent proteomic changes caused by MPXV infection, enabling us to characterize the MPX host response at the proteomic level despite the moderate cohort size of a case series, in a timely manner. We report several protein markers that correlate with disease severity in the tested cases, that classify the disease proteome, and that contrast the human host response of MPXV to that of SARS-CoV-2 infection. Because we detected a partial overlap between the MPX and COVID-19 host response proteome, we were able to explore the possibility of repurposing a COVID-19 biomarker panel for classifying MPX. We report successful classification of the MPX cases based on the proteomic biomarkers initially designed to assess severity and outcome in COVID-19 [22]. Hence, our case series study provides a biochemical characterization of the MPX host response and reveals correlation of host proteins with MPX disease severity. Furthermore, our findings suggest that there is an untapped potential in the rapid repurposing of proteomic biomarker panels across viral diseases, which would speed up response time in the event of a pandemic for the development of new therapeutics, and if needed, optimize resource allocation in health care systems.

## Materials and Methods

### Patient cohort, biosamples, and clinical data

Patients with PCR-confirmed MPXV infection were recruited in a prospective observational study on the clinical and molecular characteristics of MPX. Written informed consent for collection of clinical data and blood was obtained from all patients before inclusion. Biosampling for proteomic measurements was performed on day 1–3 after admission to the hospital. Clinical data were captured in a purpose-built database. The study was approved by the ethics committee of Charité – Universitätsmedizin Berlin (EA2/139/22). Biosamples for the cohort of patients with COVID-19 were obtained from the PaCOVID-19 study, a prospective observational cohort study on the pathophysiology of COVID-19 conducted at Charité – Universitätsmedizin Berlin [23,24]. Biosamples for the cohort of healthy controls were obtained from a clinical study including healthy volunteers [25].

### Reagents and consumables

Water was from Merck (LiChrosolv LC–MS grade; Cat# 115333), acetonitrile was from Biosolve (LC-MS grade; Cat# 012078), trypsin (sequence grade; Cat# V511X) and trypsin/LysC mix (mass-spec grade; Cat# V5072) were from Promega, 1,4-dithiothreitol (DTT; Cat# 6908.2) was from Carl Roth, urea (puriss. P.a., reag. Ph. Eur.; Cat# 33247), Tris(2-carboxyethyl) phosphine hydrochloride (TCEP; Cat# 646547), and RIPA buffer (Cat# R0278) were from Merck, ammonium bicarbonate (ABC; eluent additive for LC–MS; Cat# 40867), 2-chloroacetamide (Cat# 22788), and dimethyl sulfoxide (DMSO; Cat# 41648) were from Fluka, formic acid (LC–MS grade; eluent additive for LC–MS; Cat# 85178), PCR sealing foil sheets (Cat# AB-0626), and Pierce quantitative fluorometric peptide assays (Cat# 23290) were from Thermo Fisher Scientific, bovine serum albumin (BSA; albumin Bovine Fraction V, Very Low Endotoxin, Fatty Acid-free; Cat# 47299) was from Serva, 96-well ultrafiltration plates (AcroPrep Advance Filter Plates for Ultrafiltration, 1 ml, Omega 30K MWCO (Cat# 8165) were from PALL, 96-well LoBind plates (Cat# ER0030129512-25EA) were from Merck, stable isotopic labeled (SIL) reference peptides for discovery proteomics (PQ500 Reference Peptides) were from Biognosys.

### Sample preparation

Plasma samples were diluted 1:10 in RIPA buffer and heated at 95 °C for 10 min. After cooling to room temperature (RT), 15 µl (∼100 µg protein) of each sample were transferred to a 96-well ultrafiltration plate mounted onto a collection plate (96-well LoBind plate). Liquid was removed by centrifugation (30 min, 1800 × rcf, 20 °C). Samples were denatured and reduced in 50 µl TUA buffer (8 M urea, 20 mM ammonium bicarbonate, 5 mM TCEP) for 30 min at room temperature without shaking. Following thiol alkylation (addition of 10 µl CA buffer (50 mM 2-chloroacetamide, 20 mM ABC) and incubation in the dark at RT for 30 min), the plate was centrifuged (30 min, 1800 × rcf, 20 °C). Samples were washed twice (30 min, 1800 × rcf, 20 °C) with 100 µl 20 mM ABC. Following an additional centrifugation to remove residual liquid (60 min, 1800 × rcf, 20 °C), the filter plate was moved to a fresh collection plate. To each well 50 µl 20 mM ABC containing 1 µg of trypsin/LysC mix was added, the plate was sealed with an adhesive PCR sealing foil sheet and incubated at 37 °C for 15 h. Peptides were collected by centrifugation (30 min, 1800 × rcf, 20 °C). Following the addition of 70 µl of HPLC-grade water to each well, the plate was centrifuged once more. The collection plate was then placed in a SpeedVac and samples were evaporated to complete dryness. Peptides were reconstituted in formic acid (30 µl, 0.1% v/v). Peptide concentration was determined using the Pierce quantitative fluorometric peptide assay.

For discovery proteomics, all samples (QCs, monkeypox, COVID-19, and healthy controls) were diluted to 200 ng/µl. The stable isotopic labeled reference peptides (PQ500 Reference Peptides) stock was prepared as described in the vendor’s protocol [26]), and diluted 1:10 in 50/50 v/v ACN:H_2_O. 2 µl of diluted PQ500 stock solution were spiked into 18 µl of the 200 ng/µl sample before transfer to vials for injection. For targeted proteomics, 15 µl of pre-digested heavy labeled standards (details in [22]) were spiked into 10 µl samples (QCs, monkeypox, COVID-19, and healthy controls) and 20 µl were injected into the LC-MS system.

### Mass spectrometry

#### Discovery proteomics using Zeno SWATH MS

Tryptic digests were analyzed on a 7600 ZenoTOF mass spectrometer system (SCIEX), coupled to an ACQUITY UPLC M-Class system (Waters). 2 µl of each sample (360 ng sample + 0.02 µl PQ500, Biognosys) were loaded on a HSS T3 column (300 µm × 150 mm, 1.8 µm, Waters) heated to 35 °C, then chromatographically separated with a 20-min gradient using a flow rate of 5 µl/min [27]. A Zeno SWATH acquisition scheme with 85 variable-size windows and 11-ms accumulation time was used [28] for MS detection.

#### Targeted proteomics by multiple reaction monitoring (*plasma biomarker panel,* [22]

Tryptic digests were analyzed on a 6495C triple quadrupole mass spectrometer (Agilent) coupled to a 1290 Infinity II UHPLC system (Agilent). Prior to MS analysis, samples were chromatographically separated on an InfinityLab Poroshell 120 EC-C18 column (2.1 × 50 mm, 1.9 µm, Agilent) heated to 45 °C with a flow rate of 800 µl/min. The 6495C mass spectrometer was controlled by MassHunter Workstation software (LC–MS/MS Data Acquisition for 6400 series Triple Quadrupole, Version 10.1 (Agilent)) and was operated in positive electrospray ionization mode. Samples were analyzed in dynamic multiple reaction monitoring (MRM) mode with both quadrupoles operating at unit resolution [22].

### Data processing

#### Discovery proteomics

The Zeno SWATH raw proteomics data was processed using DIA-NN 1.8.1 beta 20 [29]. The MS2 and MS1 mass accuracies were set to 20 and 12 ppm, and the scan window to 7. For the discovery approach, we used a publicly available spectral library for human plasma [30] and replaced spectra and RT information with DIA-NN in-silico prediction. Protein inference was switched off and the match-between-runs (MBR) option was enabled.

#### Targeted proteomics

LC–MRM data were processed using MassHunter Quantitative Analysis, v10.1 (Agilent). Peptide absolute concentration (expressed in ng/ml) was determined from calibration curves, constructed with native and SIL peptide standards in surrogate matrix (40 mg/ml BSA), and manually validated. Linear regression analysis of each calibration curve was performed using custom R code (with 1/x weighting). Detailed information on transitions and matching of native peptides and internal standards can be found in [22]. Peptides with > 40% of values below the lowest or above the highest detected calibrant concentration across all samples were removed from analysis.

### Data analysis

#### Clinical data analysis

Pseudonymized clinical data were processed using JMP Pro 16 (SAS Institute).

#### Discovery proteomics data analysis

Peptide expressions were first normalized within each clinical group. Peptides with excessive missing values (> 40 % per group) were not considered in our analysis. The missing values of remaining peptides were imputed group-based using the PCA method [31]. After imputation an additional step of normalization was applied to the total set without using group information. In both cases normalization was performed with LIMMA [32] implementation of cyclic loess method [33] with option “fast” [34]. To obtain a quantitative protein data matrix, the log_2_-intensities of peptides were filtered, only peptides belonging to one protein group were kept, and then summarized into protein log intensity using the PLM method [35] implemented in the preprocessCore R package [36].

Statistical analysis of proteomics data was carried out in R using publicly available packages. Linear modeling was based on the R package LIMMA [32]. The following model was applied to each tissue dataset (log_2_(p) is the log_2_-transformed expression of a protein): log_2_(p) ∼ 0 + Class. The categorical factor Class had three levels: MPX, COVID-19, and control; reference level: control. For correlation between MPX severity (N_Skin lesions_) and protein expression, log_2_(1 + N_Lesions_ / 15) was used for linear regression.

For finding regulated features, the following criteria were applied: Significance level alpha was set to 0.015, which guaranteed for Contrast MPX vs healthy the Benjamini–Hochberg [37] false discovery rate below 5%. The log fold-change criterion was applied to guarantee that the measured signal is above the average noise level. As such we took the median residual standard deviation of linear model: log_2_(T) = median residual SD of linear modeling (= log_2_(1.35)). Functional GSEA analysis was carried out using the clusterProfiler R package [38]. For selecting the most (de)regulated pathway terms we applied filter: 3 ≤ term size ≤ 300. The data matrix and description are provided in Supp. Table 1.

#### Classifier construction and protein/peptide ranking

The classifiers were constructed using a linear support vector machine (*sklearn*.*svm*.*LinearSVC()*) as implemented in scikit-learn 1.0.2 [39] with an L1-penalty and balanced class-weights. The maximum number of iterations was increased to 10,000 to ensure convergence. As input, the log_2_-transformed quantities of the discovery proteomics and the 32 quantified peptides of the MRM panel were used, respectively.

The models were constructed and tested using a 5-fold shuffled and stratified cross-validation as implemented in *sklearn*.*model_selection*.*StratifiedKFold()*. For each iteration, 4 folds were used for training, 1 fold was used for testing the model. The data were scaled using *sklearn*.*preprocessing*.*StandardScaler()* fitted on the training data.

The AUC was calculated for the test data that were not used for training the model after all 5 iterations, resulting in one predicted value for every sample. For each iteration, the coefficients of the trained model were extracted and normalized by the maximum absolute coefficient of this iteration. For the plots, the mean and the standard deviation (error bars) of all 5 coefficients per protein/peptide were calculated and sorted according to the absolute mean. For reproducibility, the seed was fixed to 42.

#### Targeted proteomics

Significance testing of the absolute peptide concentrations and the sample type (control, MPX) was performed using Mann–Whitney U test with multiple testing correction (where indicated). Test results are provided in Supp. Table 2, p-values < 0.05 were considered significant.

## Results

### MPX patient case series and clinical presentation

A group of five patients were hospitalized at Charité University Hospital between 26^th^ and 31^st^ May 2022 for treatment of MPX, detected by PCR from cutaneous blisters. Interestingly, all patients had attended the same social event 10–14 days before developing symptoms, three of whom considered it most likely to have been infected on that occasion. We then included a 6^th^ patient with an unrelated infection history who was hospitalized on mid-June 2022, but that otherwise had a related disease history. All six patients self-identified as men having sex with men (MSM) having practiced receptive anal sexual intercourse within 14 days prior to hospitalization. The group of patients was therefore remarkably homogeneous regarding history and time course of infection, triggering our interest in a case series study. To be able to contrast the case series, we selected two control groups of age- and sex-matched healthy controls (the case series cohort age ranged from 26 to 49 years) and patients with moderate COVID-19 (WHO grade 3, i.e., hospitalized without the need of supplemental oxygen therapy), respectively (Methods).

Overall, MPX patients exhibited mild to moderate symptoms. Prodromes included fever, myalgia, and fatigue, and had already subsided in all patients by the time of admission to the hospital. The number of MPX skin lesions ranged from 5 to 36 and there were no clinical or laboratory signs of organ dysfunction. In all patients, the chief complaint and cause of hospitalization was severe anal or perianal pain requiring systemic analgesics in addition to topical treatment. Samples for proteome measurements were taken at a median of 8 days after symptom onset. Comorbidities included HIV (n = 2, both well controlled on antiretroviral therapy), other STIs (n = 1), and hepatitis C (n = 1). Patients were discharged with alleviated symptoms after 3–6 days. A summary of clinical characteristics is given in **Table 1**.

**Table 1:** Patient characteristics.

|  | MPXV cases (n = 6) |  |
| --- | --- | --- |
| male, n (%) | 6 | 100% |
| age, years | 31 | 27–41; 26–49 |
| BMI, kg/m<sup>2</sup> | 22.0 | 19.6–23.4; 17.6–25.1 |
| comorbidities, n (%) | 3 | 50% |
| HIV, n (%) | 2 | 33% |
| hepatitis C, n (%) | 1 | 17% |
| other STIs*, n (%) | 1 | 17% |
| Δ symptom onset to sample, days | 8 | 5–14; 5–17 |
| Δ PCR to sample, days | 3.5 | 1.5–5; 0–5 |
| fever, n (%) | 6 | 100% |
| number of lesions | 9 | 5–20; 5–36 |
| duration of hospital stay, days | 3.5 | 3–5; 3–6 |
| C-reactive protein at admission, mg/l | 20.0 | 10.4–57.9; 8.7–120.8 |
| leukocytes at admission, per nl | 9.7 | 8.3–11.7; 8.1–12.9 |
| lymphocytes at admission, per nl | 3.1 | 1.6–3.7; 1.4–3.8 |
| lactate dehydrogenase, U/l | 214 | 203–273; 181–381 |
| Data are presented as median, IQR; range unless otherwise specified. BMI: body mass index; HIV: human immunodeficiency virus infection; STI: sexually transmitted infection.<br>* Other STIs: co-infection with *Neisseria gonorrhoeae*, *Ureaplasma*, and *Mycoplasma hominis*. |  |  |

The partial sequence of the genome of the MPXV isolate obtained from one of the patients was determined and is available on GenBank (ON813251.2).

To gain maximum information from the case series cohort, we assembled two control cohorts. The first consisted of 15 age- and sex-matched healthy volunteers (Supp. Table 3). Ten patients with SARS-CoV-2 infection, hospitalized due to moderate COVID-19 (grade 3 on the 8-point WHO ordinal scale, i.e., without the need for supplemental oxygen therapy), constituted the second control group. Their proteomes were measured within the same batch on our MS platforms, but had also been analyzed by us as part of a previous study [15] (Supp. Table 3)

### A plasma proteomic signature of MPXV infection

Because of the moderate size of the case series study, we focused on obtaining maximally precise proteomic measurements, and contrasted against both control groups. For obtaining precise proteomic measurements, we prepared tryptic digests from the MPX cases, matched healthy controls, and moderate COVID-19 patients using a plasma proteomics sample-preparation platform optimized for high precision. Moreover, we included a broad panel of stable-isotope-labeled internal standards (PQ500, Biognosys). The tryptic digests obtained were then recorded using an online coupling of microflow chromatography and Zeno SWATH DIA, the latest generation of DIA proteomic technology [28]. Indeed, to our knowledge, the present study represents the first biomedical application of Zeno SWATH MS. After data were recorded as a single batch, raw data were processed with DIA-NN [40], and data were post-processed to detect differentially concentrated proteins as well as the enrichment of pathway terms using pathway definitions from REACTOME [41]. A workflow diagram of the procedures is provided **(Fig. 1a)**.

**Figure 1.**
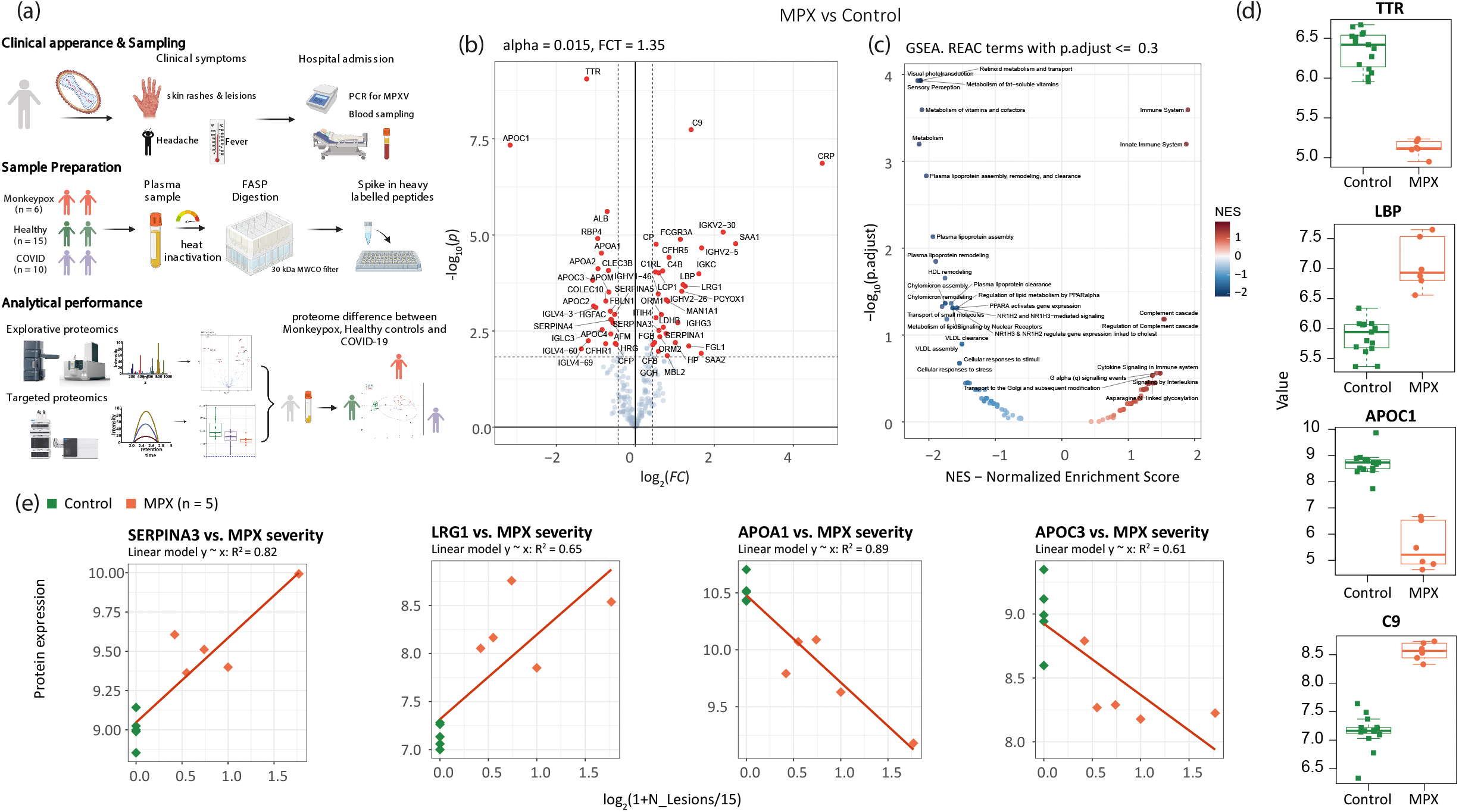
The human host response to monkeypox virus infection determined at the level of the plasma proteome. **a)** Schematic overview of the workflow using discovery proteomics (Zeno SWATH MS [28]) in parallel to a targeted proteomic assay that quantifies COVID-19 severity biomarkers ([22]) to characterize the plasma proteome in an MPX case series, and comparison the proteomes to those of healthy volunteers and COVID-19 patients. **b)** Volcano plot of contrast MPX vs healthy controls; α <= 0.015 and |logFC| >= 1.35 were used for selection of regulated proteins. **c)** Gene set analysis (GSEA) of REACTOME [41] terms enrichment for contrast MPX vs control. Y-axis shows –log_10_ of adjusted p-value (fdr) for Normalized Enrichment Score (x-axis) for each term. Terms with fdr <= 0.3 are labeled. **d)** Boxplots illustrating key proteins that differ between patients with MPX and controls. **e)** Correlation between MPX severity (N_Skin lesions_) and protein expression (y-axis). One MPX patient had an unclear additional skin condition (not a pure case of MPX) and therefore was excluded from the regression analysis that compares the number of skin lesions with the proteome; however, the proteome of this patient was largely in agreement with those of the other MPX cases (**Supp. Fig. 3**). As a measure of MPX severity, the log_2_(1 + N_Lesions_ / 15) was used. Here N_Lesions_ is the number of lesions. R^2^ shows squared correlation coefficient. MPX patients are colored orange, control patients green.

Considering the relatively mild severity of clinical symptoms and skin manifestation, the data revealed a substantial proteomic response to MPXV infection within the abundant ‘functional fraction’ of the plasma proteome. This proteome fraction constitutes more than 99.9% of the plasma proteomic mass and is composed of around 300 proteins, most of which directly function in the plasma [42]. As 200– 300 of them are consistently quantified using high-throughput proteomics in neat plasma [12], and because this fraction contains more than 50 typical protein biomarkers [15] that capture host physiological parameters [43], thia functional fraction of the plasma proteome is of special interest for the development of clinical assays [22]. After pre-processing, 226 of the highly abundant proteins were found consistently quantified in the neat plasma sample. We detected low within-group coefficients of variation, below 25% for MPX and control, and about 34% for COVID-19 cases, indicating a high quantitative precision of the measurements, but also the presence of a biological signal (**Supp. Fig. 1c**). Indeed, we found 56 of the major plasma proteins to be differentially abundant in MPX patients compared to healthy controls. 24 of these were lower concentrated in MPX, and 32 detected at a higher concentration (**Fig. 1b**). The nature of the affected proteins indicated the molecular processes affected by MPX, as revealed by an enrichment analysis. For example, we see “Immune system” and “Regulation of complement cascade” mostly enriched among upregulated pathways. Among downregulated pathways, “Plasma lipoprotein assembly” and “Metabolism of fat-soluble vitamins” are enriched (**Fig. 1c**).

At the level of individual proteins, the greatest differences between cases and controls were found in proteins associated with the acute phase response. These included significantly lower levels of the negative acute phase proteins TTR, ALB, and RBP4, as well as higher levels of acute phase proteins CRP, SAA1, SERPINA3, LBP, CP, and LRG1. Of note, various proteins involved in hepatic lipid metabolism and nutrient transport (APOA1, APOA2, APOC1, APOC2, APOC3) were lower in MPX patients than in controls, a known but not fully understood phenomenon also observed in other infections [44] (**Fig. 1d**). Compared to controls, MPX patients exhibited a significantly higher level of complement component 9, the main element of the channel part of the membrane attack complex. Also, TTR in combination with the differentially expressed apolipoproteins is noteworthy, as it is a marker for malnutrition [45] and we recently found it as a rapid responder in a caloric-restriction experiment conducted with healthy volunteers [43]. We first speculated that acute MPX could result in a reduced caloric intake in affected patients. However, this picture was not confirmed by the clinical records of our patients, indicating that also TTR is part of the host response. Furthermore, we did not observe an influence of the concomitant conditions such as HIV on the plasma proteomes, which is reasonable as all patients with HIV had immunologically well-controlled infections with suppressed viral load (**Supp. Fig. 2**).

Next, we tested whether there is a relationship between the proteomic response and the number of skin lesions observed in our patients, determined as a proxy of disease severity. Several peptides showed a statistically robust correlation with the number of lesions, including the upregulated acute phase proteins SERPINA3, SAA1, and LRG1, as well as the downregulated apolipoproteins APOA1, APOA2, and APOC3 (**Fig. 1e**). In particular LRG1, an upstream modifier of TGF-beta signaling, is being increasingly recognized as an important contributor to disease pathogenesis and hence as a potential therapeutic target in a range of inflammatory conditions [46]. Despite the moderate size of the case series, our data suggests a consistent proteomic response in MPX cases that reflects the extent of skin manifestation and disease severity in MPX.

### Relationship and intersection of the acute phase proteomic responses of MPX and COVID-19

The plasma proteome has similarly been shown to distinguish between different degrees of disease severity in other viral infections, including ebola [47] and COVID-19 [15–17,19,20]. To investigate to which degree this classification is due to a similar or divergent set of protein markers, we compared the MPX proteome response to that of an age- and sex-matched group of patients with moderately symptomatic COVID-19 (hospitalized, but without need of supplemental oxygen). The proteome obtained for these two patient groups revealed both an overlap in some response proteins, and differences between the host responses against the two viral pathogens in other proteins. A simple hierarchical clustering based on Ward’s agglomeration of Euclidean distances clearly separated healthy controls from MPX and COVID-19 cases (**Fig. 2a**), and a protein expression analysis revealed differentially expressed proteins that are common between both diseases, but also those that differentiate the two infections from each other (**Fig. 2b** (central part of the cloud), full-scale figure in **Supp. Fig. 4a**). Consistently, a principal component analysis (PCA) separated both patient groups (and controls), indicating that despite an overlap in several factors, the proteomes are discriminatory between MPX and COVID-19 (**Fig. 2c**).

**Figure 2.**
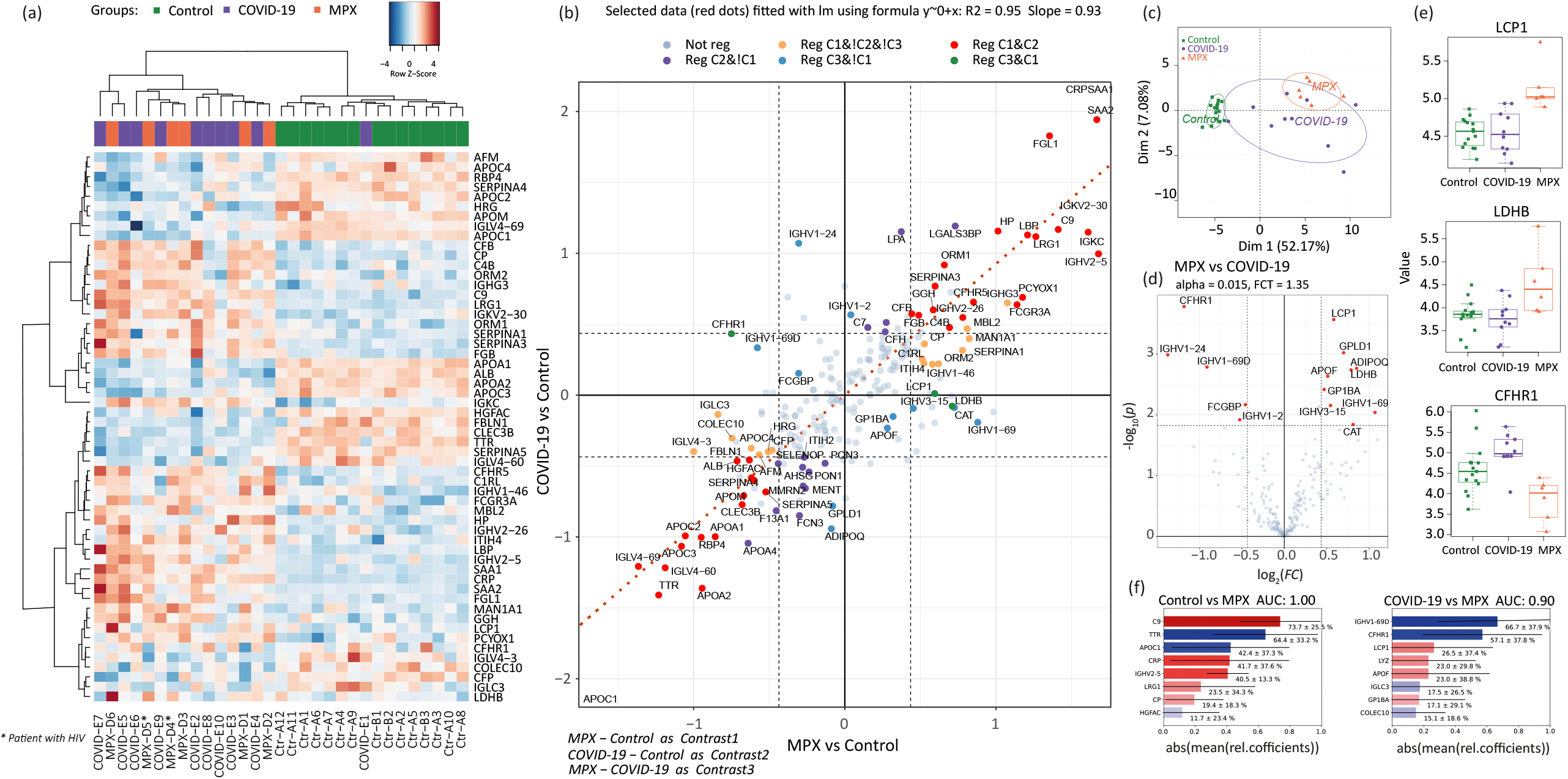
Differences and similarities between the plasma proteome upon infection with MPXV and SARS-CoV-2. **a)** Heatmap displaying hierarchical clustering using differentially regulated proteins between patients with MPX, COVID-19, and controls. **b)** Scatterplot of log fold-change (logFC) for contrast MPX vs control (C1, x-axis) and logFC for contrast COVID-19 vs control (C2, y-axis). Only the central part of the cloud is shown here. Three truncated dots (APOC1, CRP, and SAA1) are shown in the lower left and upper right corner. A full-scale figure is presented in **Supp. Fig. 4a**). Differentially regulated (‘Reg’) proteins are color coded, with the red color corresponding to 37 proteins regulated in both MPX vs control (C1) and COVID-19 vs control (C2), the orange color corresponding to proteins specific for MPX vs control (C1) only (16 proteins), and the green color corresponding to proteins regulated both in MPX vs control (C1) and MPX vs COVID-19 (C3) (3 proteins). There are no intersections between COVID-19 vs control (C2) and MPX vs COVID-19 (C3). The blue color corresponds to proteins regulated in MPX vs COVID-19 (C3), but not in MPX vs control (C1) (11 proteins), and the pink color to proteins regulated in COVID-19 vs control (C2) only (19 proteins). The red dotted line shows a linear regression through the red dots, i.e., proteins regulated in MPX vs control (C1) and COVID-19 vs control (C2). Note that orange and pink points have the same direction of regulation in both MPXV vs control (C1) and COVID-19 vs control (C2). Only green and blue dots (except three proteins: ADIPOQ, GPLD1, and IGHV1-2) have opposite directions in C1 and in C2. **c)** Post hoc PCA score plot using proteins shown in (a). **d)** Volcano plot showing differentially regulated proteins of patients with MPX and COVID-19; α <= 0.015 and |logFC| >= 1.35 were used for selection of regulated proteins. **e)** Boxplots illustrating key proteins that differ between patients with MPX and COVID-19. **f)** Top 8 proteins of an SVM-trained model discriminating between healthy controls and MPX cases (top) or COVID-19 and MPX cases (bottom). Means of the relative coefficients over a 5-fold cross-validation are shown. Error bars denote the standard deviations. Red denotes positive, blue denotes negative coefficients. The AUC was calculated based on withheld samples that were not used for training the model.

Contrasting the signatures at the protein level revealed that of the 56 proteins differentially expressed in MPX cases compared to healthy controls, 37 are also differentially expressed in COVID-19 patients with the same direction of regulation (**Supp. Fig. 4a**, Venn diagram). These include 12 proteins of the acute phase response such as SAA1 and LBP, and 12 proteins involved in coagulation, including FGB and SERPINA4, all of which have been found to be differentially expressed depending on COVID-19 disease severity.

Furthermore, we found 19 proteins that were differentially abundant in MPX but not in COVID-19. For instance, LCP1 and LDHB were found to be only upregulated in MPX (**Fig 2 d, e**). LCP1 is interesting, because as L-plastin, it has been associated with membrane dynamics and the cytoskeleton and is an early tumor marker in kidney cancer [48,49]. Another protein that triggered our attention was CFHR1, an inhibitor of the terminal pathway of the complement cascade, which was downregulated in MPX but was upregulated in COVID-19, where it is a marker of disease severity [15–17]. Indeed, hyperactivation of the complement system has been shown as a key feature for the pathophysiology of COVID-19 [50], but according to our proteome data, is less important in MPX.

In the next step, we tested a cross-validated classifier to distinguish between MPX cases and healthy controls, as well as between MPX and COVID-19 cases on the basis of their proteomes. For both cases we achieved a differentiation with a high accuracy (**Fig. 2f**). Encouragingly, the top-ranked differentiators identified by the machine-learning regression were also among the most differentially expressed proteins, like C9 and TTR for differentiating MPX cases and healthy individuals, or CHFR1 or LCP1 for differentiating MPX from COVID-19, respectively (**Fig. 2f**).

Hence, our data provide a differentiated picture of the acute proteomic response that follows the two viral infections. On the one hand, we describe various acute phase proteins responding to both COVID-19 and MPX; on the other hand, both viral infections exhibit distinct proteomic response patterns, for instance concerning the activation of the complement system. Hence, proteomics was effective in obtaining valuable insights even from case series studies.

### Potential to repurpose proteomic assays to rapidly respond to emerging viral infections

Emerging pathogens with pandemic potential require fast responses, and an attractive possibility to achieve that is in the repurposing of existing procedures, diagnostics, and therapies, whenever possible. Prognostic biomarker panel assays were discussed during the COVID-19 pandemic for the monitoring of clinical trials, for supporting clinical decisions, and for their potential to support the navigation through difficult triaging situations [11,22,51]. Due to the partial overlap between the COVID-19 and MPX host responses, we speculated that it might be possible to repurpose COVID-19 biomarker panel tests for MPX. We recently demonstrated the translational potential of plasma proteomics for applicability in clinical practice through the transfer of protein marker candidates which had been identified by discovery proteomics in COVID-19 into a routinely applicable targeted protein panel assay. The assay absolutely quantifies up to 50 peptides derived from 30 COVID-19-related plasma biomarker proteins and captured hallmarks of COVID-19 in a multi-cohort observational study conducted using routine-lab-compatible high-flow chromatography and LC-MRM acquisition [22]. The LC–MRM assay consistently quantifies 32 of the peptides in plasma samples from MPX cases, controls, and in COVID-19 patient samples. Despite the assay being developed to quantify COVID-19 severity, a PCA on the peptides quantified separated also MPX patient samples from controls (**Fig. 3a**). Moreover, a hierarchical clustering of the protein quantities that differed between healthy controls and MPX cases classified the disease samples (**Fig. 3b**). This separation was driven by differential plasma levels of several proteins involved in the inflammatory and immune-mediated host response, e.g., increased levels of SERPINA3 and LYZ, or decreased levels of TF, TTR, HRG, PGLYRP2, and APOA1 (**Fig. 3c**).

**Figure 3.**
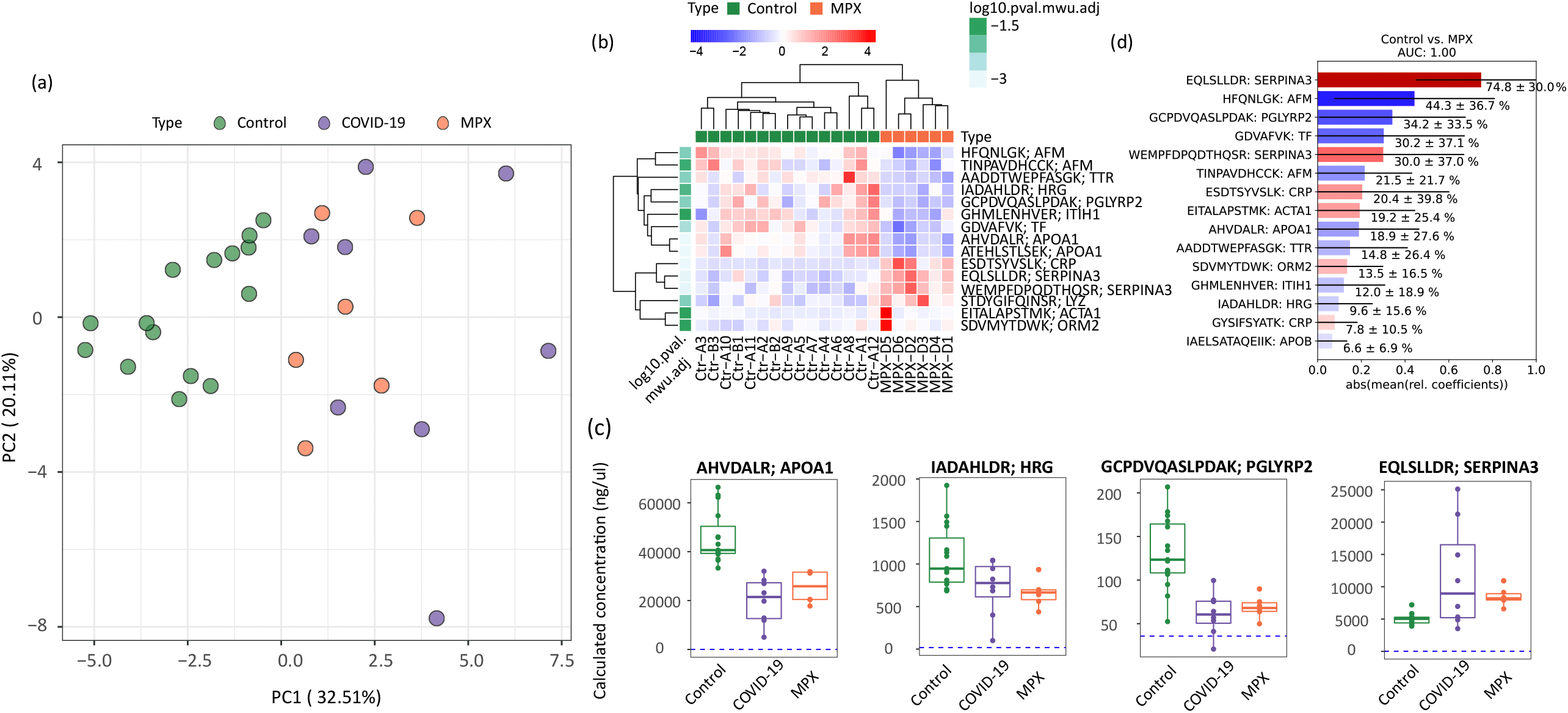
A targeted, multi-protein panel assay developed for COVID-19 infection discriminates patients with MPX from controls. **a)** Principal component analysis (PCA) of controls, patients with MPX, and COVID-19 with 32 peptides absolutely quantified in all samples. **b)** Heatmap displaying hierarchical clustering using differentially regulated proteins between patients with MPX and controls; p < 0.05 with Mann–Whitney U test with FDR-based multiple testing correction. **c)** Boxplots illustrating key proteins that differ between patients with MPX and controls, and COVID-19. Dashed blue lines indicate the lowest detected peptide concentration from calibration curves. **d)** Top 15 peptides of an SVM-trained model discriminating between healthy controls and MPX cases. Means of the relative coefficients over a 5-fold cross-validation are shown. Error bars denote the standard deviations. Red denotes positive, blue denotes negative coefficients. The AUC was calculated based on withheld samples that were not used for training the model.

Based on this proteomics data, we constructed a classifier, that distinguished between MPX cases and healthy controls (**Fig. 3d)**. The most important features of the classifier (e.g., SERPINA3, AFM, PGLYRP2, and TF) did overlap with the differentially concentrated proteins in the disease plasma samples.

## Discussion

As case numbers rise rapidly, the current knowledge gap on the molecular etiology of MPX—a disease that has been known in central Africa for more than 50 years—becomes ever more apparent and calls urgently for a better understanding of this disease. In this context, a case series of individuals with similar demographics, timing, and course of disease who likely contracted the infection at the same social event caught our attention. Usually, the host response to a viral pathogen would be investigated in larger cohorts. However, considering urgently needed data and the parallel disease history of our case series, we speculated that because of the homogeneous and representative nature of cohort considering the current outbreak, even a low number of individuals may provide a clear proteomic signal, allowing us to provide a timely assessment of the host response to MPXV infection.

Indeed, analyzing the host response of the MPX patients at the proteome level provided a surprisingly clear picture, even in this small cohort, especially when comparing the proteomes to age- and sex-matched healthy individuals or patients with severity-matched, moderate COVID-19. Our dataset showed increased levels of specific acute phase proteins and overall lower nutritional response proteins such as TTR and apolipoproteins in MPX when compared to healthy controls. However, key pathways altered in COVID-19, including the complement and coagulation systems, were affected to a much lesser extent. The proteomic response described in our study therefore reflects the different pathophysiology connected with MPXV and SARS-CoV-2 as well as the mild to moderate disease severity in MPX observed in the current outbreak so far [9,10]. Additional cohort studies will be required to validate our results in the broader context. Reassuringly however, most proteins identified by our non-targeted proteomic technique to be differentially abundant in MPX, have a known biological role in the acute phase response to viral infections. The correlation of numerous of the inflammatory proteins with disease severity gives additional and orthogonal confidence in our results.

We identified several peptides that showed a statistically robust correlation with disease severity as determined by the number of skin lesions. Organ dysfunction and severe disease have so far only sporadically been reported in the current outbreak in Europe and the US, and there were no fatal cases reported yet [10]. MPX is however known to cause severe and lethal disease in endemic regions in Africa, with reported case fatality of up to 10% [4]. Drawing from our previous experience and based on the signature of the MPX human host response in discovery proteomics in the present study, we were able to apply a biomarker panel designed to classify patients with COVID-19 in routine laboratories

[22] to this different viral disease. The biomarker panel captured hallmarks of MPXV infection and facilitated a classification of patients with MPX and healthy controls in our sample set using an SVM model. The attractiveness of MRM panel assays is that they can be implemented in clinical workflows, are of low cost per sample. The respective peptides identified as potential severity markers could be of help for severity classification of MPX in endemic regions and possibly help to elucidate the pathophysiological differences between the Central African and West African clade of MPXV in the future. Our case series was too small to determine if the biomarker panel can be used to predict disease features, e.g., time to recovery, or to discriminate the effectiveness of therapeutic options. However, the robust correlation of the proteomic response with the number of skin lesions suggests that a predictive application of proteomics is possible for MPX, and we hope our case series study stimulates respective cohort studies in the near future.

We believe our study demonstrates two essential aspects which are important for pandemic preparedness. First, our study exemplifies that when time is of the essence, proteomics can deliver valuable information on the molecular disease etiology of a moderate number of affected individuals, at least when their disease history is homogeneous and/or representative as in our case series study. Our results therefore imply that plasma proteomics might be particularly valuable for rare and neglected diseases, where proteomics may become an increasingly attractive toolkit for systemic analyses, despite limited case numbers. Indeed, given that symptoms were relatively mild, the proteomic host response to MPXV was distinct, with about one quarter of the highly abundant functional fraction of the plasma proteome changing. Second, our study shows that a biomarker panel assay developed for COVID-19 can be repurposed for another viral infection, as the partial overlap of the response proteome between MPX and COVID-19 was sufficient to achieve disease classification. The repurposing of dedicated panel assays to (re-)emerging pathogens could accelerate the response to a rapidly developing epidemic. In practice, our findings imply that a general plasma proteomics panel assay could potentially capture a pan-pathogen host response, merely requiring adaptation of the data analysis procedure (i.e., the classifiers and predictive models) to render the test applicable to a new pathogen. To our knowledge, the repurposing of panel assays for assessing disease severity, for clinical trial monitoring, or for predicting future disease courses has not been explored previously but could prove a helpful tool in mounting a rapid response to emerging diseases.

## Data Availability

Internal patient IDs were replaced at random within groups. Data matrix for discovery proteomics is available in (Supp. Table 1). Proteomic raw data will be made available on Pride (https://www.ebi.ac.uk/pride/) upon publication of the study.

## Acknowledgements

Fig. 1a was made using BioRender (Biorender.com). We thank Vadim Demichev, Eva Tranter, Gisèle Godzick-Njomgang, Daniel Wendisch, Linda Jürgens, and Frieder Pfäfflin for their valuable input and help in collecting biosamples, Hezi Tenenboim for proofreading our manuscript. This research was funded in part by the Berlin Institute of Health (BIH), Wellcome Trust (IA 200829/Z/16/Z to M.R), by the Ministry of Education and Research (BMBF), as part of the National Research Node ‘Mass spectrometry in Systems Medicine (MSCoresys), under grant agreement 031L0220 (to M.R). We further acknowledge funding by Deutsche Forschungsgemeinschaft (DFG) supporting Z.W.’s PhD studies as part of the TRR 186.

## Data availability

Internal patient IDs were changed at random within groups. Data matrix for discovery proteomics is available in (**Supp. Table 1**). Proteomic raw data will be made available on Pride (https://www.ebi.ac.uk/pride/) upon publication of the study.

