## Supplementary tables for "The human host response to monkeypox infection: a proteomic case series study": Supp. Table 3 - control cohort characteristics.docx

|  | **healthy controls (n=15)** | | **COVID-19 controls (n=10)** | |
| --- | --- | --- | --- | --- |
| **male, n (%)** | 15 | 100 % | 10 | 100 % |
| **age, years** | 31 | 26-45; 23-50 | 39.5 | 25-47.5; 21-50 |
| **BMI, kg/m^2^** | 23.1 | 21.2-24.9; 18.6-26.5 | 26.8 | 21.1-31.1; 20.0-39.2 |
| **Supplementary Table 3**: Control cohort characteristics. COVID-19 controls were hospitalized without need of supplemental oxygen therapy.  Data are presented as median, IQR; range unless otherwise specified. BMI: body mass index | | | | |
