## Supplementary figures for "The human host response to monkeypox infection: a proteomic case series study"

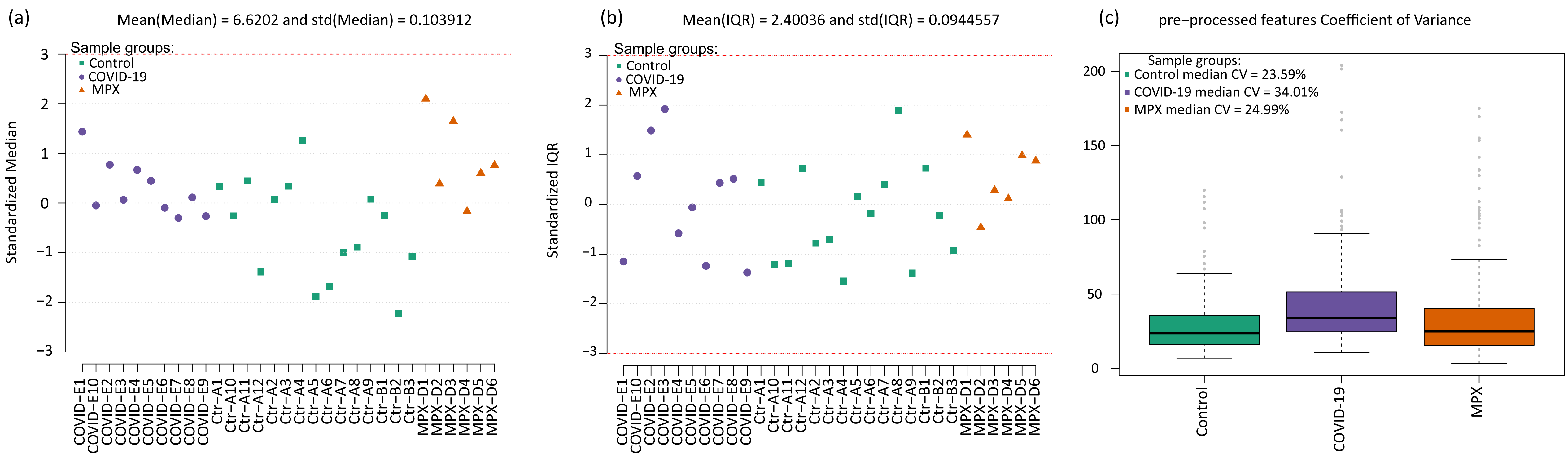


Supplementary Figure 1. Quality control charts (left and middle panels) and within group coefficient of variation (right panel)

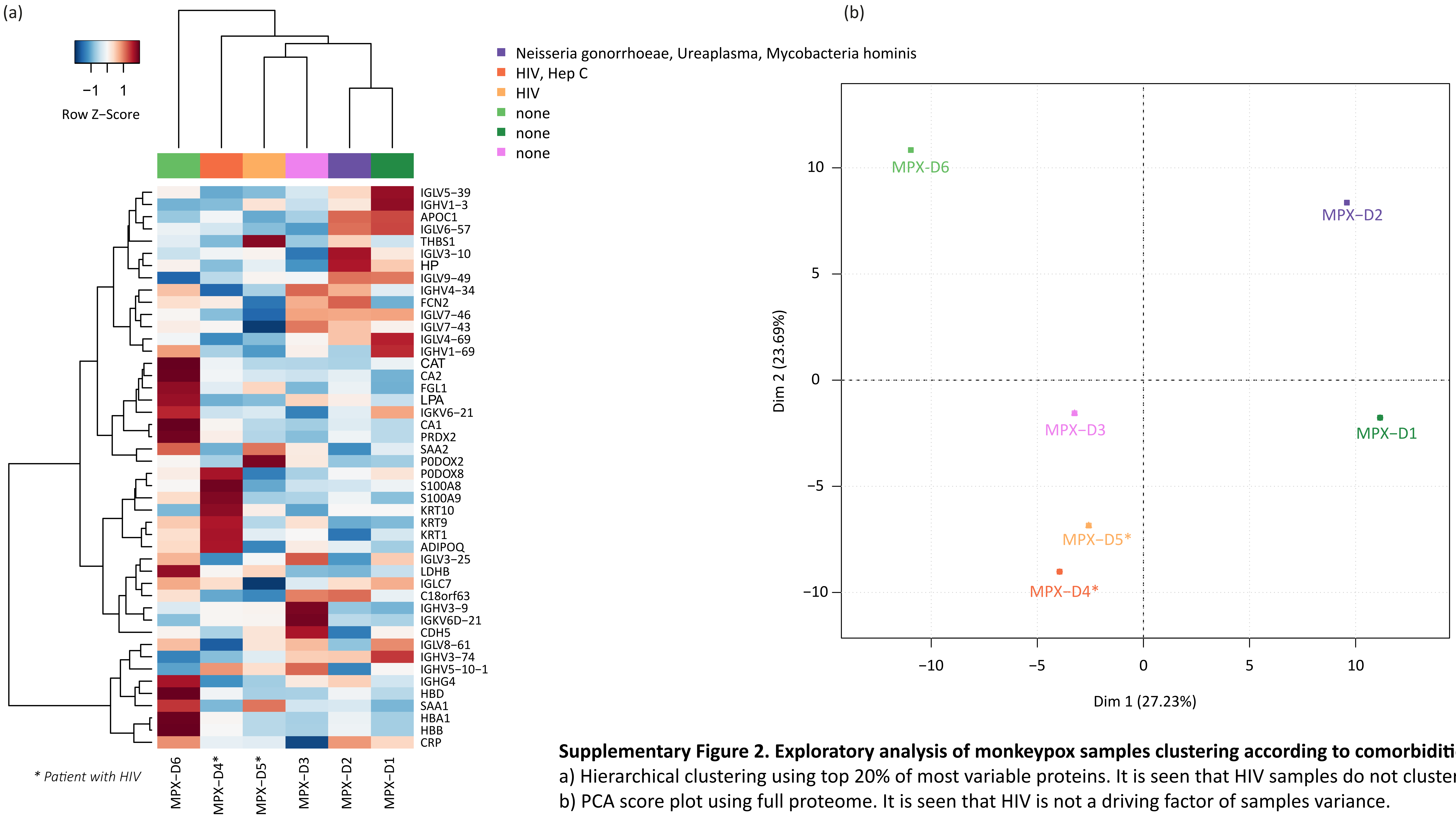


**Supplementary Figure 2. Exploratory analysis of monkeypox samples clustering according to comorbidities.**
a) Hierarchical clustering using top 20% of most variable proteins. It is seen that HIV samples do not cluster together.
b) PCA score plot using full proteome. It is seen that HIV is not a driving factor of samples variance.
Its largest contribution is into the second principal component.

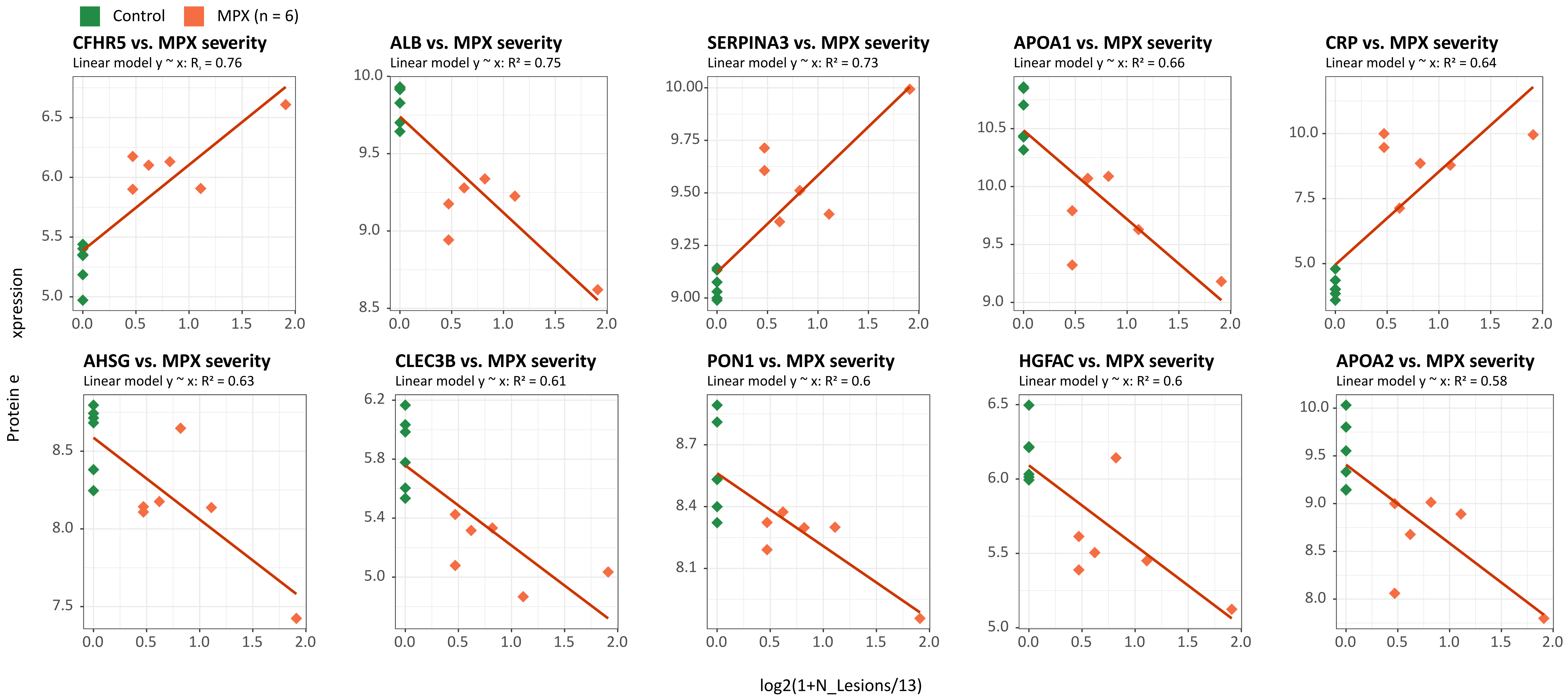


**Supplementary Figure 3. Correlation between MPX (n = 6) severity (x-axis) and protein expression (y-axis) .**

As a measure of MPX severity the $\log_2(1+N_{Lesions}/13)$ was used. Here $N_{Lesions}$ is the number of lesions.
R2 shows squared correlation coefficient. 6 MPX patients are coloured orange, age matched Control patients are coloured green.
Fitted linear dependences of protein expression vs disease severity (here $\log_2$(1+N_Lesion/Nmean), Nmean = 13) is shown as red line.
Top ten proteins with highest $R^2$ are shown.

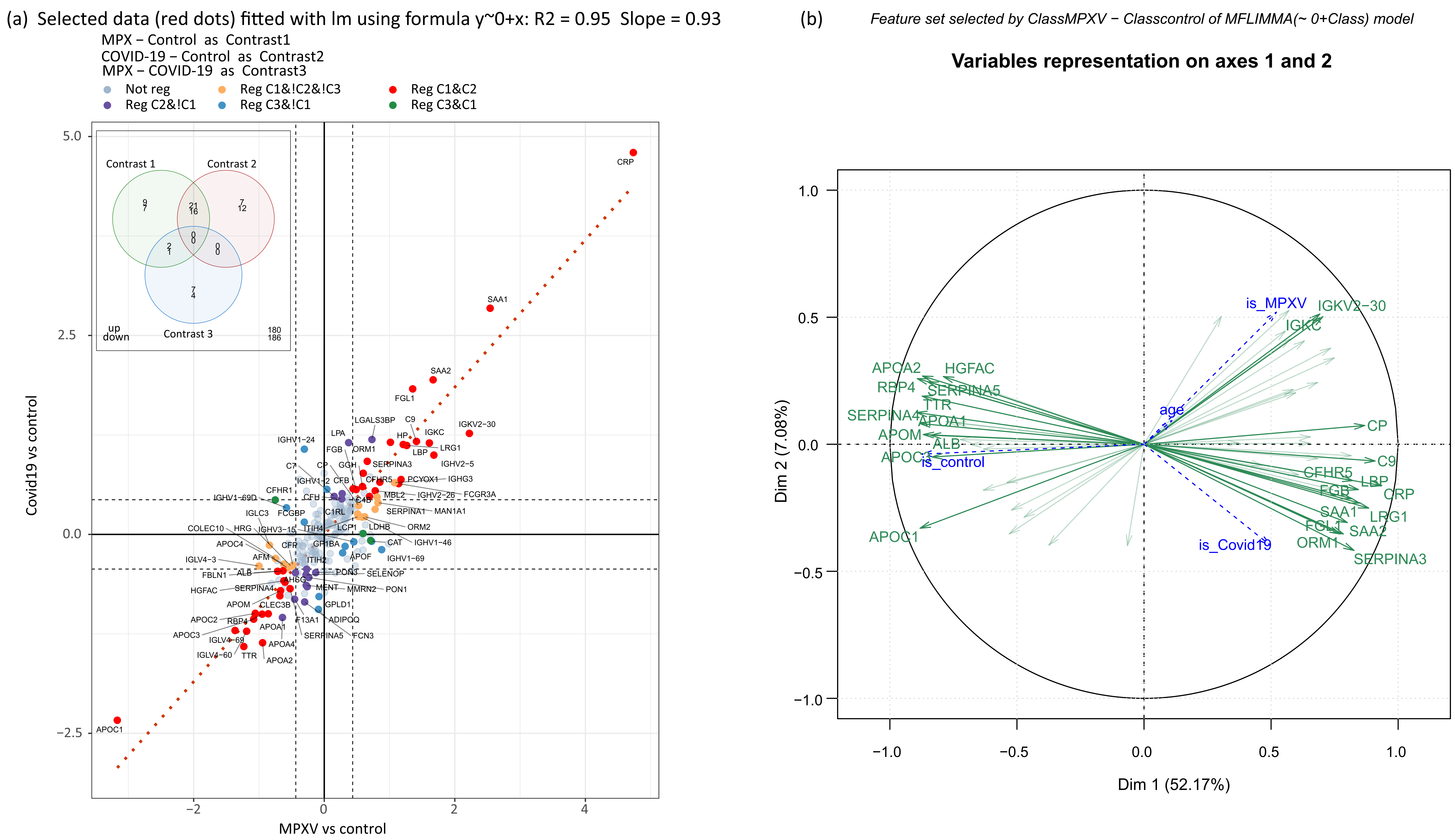
(a) Selected data (red dots) fitted with lm using formula y~0+x: R2 = 0.95 Slope = 0.93
MPX – Control as Contrast1
COVID-19 – Control as Contrast2
MPX – COVID-19 as Contrast3
Not reg
Reg C1&!C2&!C3
Reg C1&C2
Reg C2&!C1
Reg C3&!C1
Reg C3&C1
Contrast 1
Contrast 2
Contrast 3
up
down
Covid19 vs control
MPXV vs control
CRP
SAA1
SAA2
FGL1
APOC1
(b) Feature set selected by ClassMPXV − Classcontrol of MFLIMMA(~ 0+Class) model
Variables representation on axes 1 and 2
Dim 1 (52.17%)
Dim 2 (7.08%)
is_MPXV
is_Covid19
is_control
age
IGKV2−30
IGKC
APOA2
HGFAC
RBP4
SERPINA5
TTR
SERPINA4
APOA1
APOM
ALB
APOC1
CP
C9
CFHR5
LBP
FGB
CRP
SAA1
LRG1
FGL1
SAA2
ORM1
SERPINA3

**Supplementary Figure 4.** a) Scatterplot of logFC for contrast MPX vs Control (C1, x-axis) and logFC for contrast COVID-19 vs Control (C2, y- axis).
Regulated proteins are colour coded. Red colour corresponds to (37) proteins regulated in both contrasts C1 and C2.
There are no intersections between contrasts C2 and C3 (MPX vs. COVID-19). Orange group is for proteins specific for contrast C1 only (16 proteins).
Green colour is for proteins regulated both in contrast C1 and in contrast C3 (3 proteins). Blue colour is for proteins regulated in C3, but not in C1 (11 proteins).
And the pink colour is for proteins regulated in C2 only (19 proteins). Red dotted line is linear regression through red dots, i.e. proteins regulated in C1 and C2.
Note that orange and pink points have the same direction of regulation in both contrasts, C1 and C2.
Only green and blue ones (except three proteins, ADIPOQ, GPLD1 and IGHV1-2) have opposite directions in C1 and in C2.
Insert shows Venn diagram for intersection of up- and downregulated proteins between three contrasts.
b) PCA loading plot based on proteins regulated in contrast MPX vs Control. It can be seen that despite the proteins were selected for MPX vs Control,
the first principal component is in the direction of MPX + COVID-19 vs. Control and so the most of regulated proteins are.
